# Liver fat accumulation contributes to discordant genetic risk between coronary artery disease and type 2 diabetes

**DOI:** 10.64898/2026.08.24.26361276

**Authors:** Xiyun Jiang, Nick Hirschmüller, Henry J. Taylor, Mayank Dalakoti, Elise J. Needham, Martin Kelemen, Tao Jiang, Scott C. Ritchie, Antonio Vidal-Puig, Adam S. Butterworth, Samuel A. Lambert

**Affiliations:** British Heart Foundation Cardiovascular Epidemiology Unit, Department of Public Health and Primary Care, University of Cambridge, Cambridge, UK; Victor Phillip Dahdaleh Heart and Lung Research Institute, University of Cambridge, Cambridge, UK; Health Data Research UK Cambridge, Wellcome Genome Campus and University of Cambridge, Cambridge, UK; Center for Precision Health Research, National Human Genome Research Institute, National Institutes of Health, Bethesda, MD, USA; British Heart Foundation Centre of Research Excellence, University of Cambridge, Cambridge, UK; Cambridge Baker Systems Genomics Initiative, Department of Public Health and Primary Care, University of Cambridge, Cambridge, UK; MRC Institute of Metabolic Science, Metabolic Research Laboratories, University of Cambridge, Cambridge, UK; Centro de Investigacion Principe Felipe, Valencia, Spain

## Abstract

**Background:** Type 2 diabetes (T2D) and coronary artery disease (CAD) frequently co-occur, yet the biological pathways that jointly determine risk remain incompletely understood. Most genetic studies have examined shared risk from a single-disease perspective, limiting insight into the mechanisms that generate discordant risk between conditions.

**Methods:** We applied PLACO to multi-ancestry GWAS data of T2D and CAD to identify shared loci, prioritising shared causal signals using colocalisation. Shared variants were clustered by their associations with 77 cardiometabolic traits, and cluster-specific genetic risk scores (GRS) were tested for association with 17 clinical biomarkers and 1,254 binary outcomes in 378,772 UK Biobank (UKB) participants. Two-sample Mendelian randomisation (MR) was used to test the causal role of liver fat.

**Results:** We identified 149 loci shared between T2D and CAD; most novel loci had discordant effects (35 of 42), in contrast to the predominantly concordant signals reported previously. Clustering 187 independent shared variants revealed seven mechanistic clusters, three of them centred on liver fat and defined by discordant T2D–CAD effects. Enrichment analyses and cluster-GRS associations in UKB highlight associations between higher liver fat and T2D risk with a cardioprotective lipid profile and reduced CAD risk. Genetically higher liver fat increased T2D risk but lowered CAD risk in MR analyses; partitioning liver fat instruments by their effect on ApoB-containing lipoproteins indicates that the CAD effects are determined more by effects of circulating ApoB rather than liver fat itself.

**Conclusions:** Liver fat largely sets the direction of T2D risk, whereas the fate of that lipid, retained in the liver with low circulating ApoB or exported as ApoB-containing lipoproteins, sets the direction of CAD risk. This liver-centric partitioning provides a mechanistic framework for the discordant cardiometabolic effects of hepatic lipid and lipid-lowering pathways, with implications for precision prevention.

## Introduction

Type 2 diabetes (T2D) is a strong risk factor and a comorbidity of coronary artery disease (CAD), conferring a 2-fold risk increase (1,2). Around 30% of all T2D patients also have CAD(3) and T2D is responsible for around 11% of all cardiovascular deaths(4). Despite the frequent co-occurrence of T2D and CAD, there is incomplete knowledge of the shared mechanisms and risk factors. Recent studies suggest an interesting interplay of molecular mechanisms between T2D and CAD that warrants further investigation. For example, observational studies and clinical trials found that different classes of T2D medications exert varying effects on the risk of CAD, and vice versa. Most medicines used to manage T2D (e.g. GLP-1 agonists(5–10) and SGLT2 inhibitors(11)), are associated with decreased risk of CAD, at least in part independent of their glucose-lowering effect; however, sulfonylureas are generally considered neutral for CAD(12), although there have been studies reporting mixed directions of effects (13–16). For CAD medications, high intensity statin treatment is associated with an increased risk of T2D (17,18), whereas findings across studies have been conflicting for other classes of medicines (e.g. PCSK9 inhibitors(19,20) and NPC1L1 inhibitors(21,22)). Gaining insight into concordant and discordant shared pathways could inform drug discovery, repurposing and precision medicine where therapeutics may improve the outcome for one disease but confer risk to another. With the availability of large-scale genome-wide association studies (GWAS) for T2D and CAD, and the growing availability of multi-omics data, there are now new opportunities to gain a deeper understanding of shared aetiology.

Although several studies(23–28) have investigated the shared aetiology between T2D and CAD, they were limited in several ways. For example, a study identified 175 shared loci in East Asians and 602 loci in Europeans(28); however, the majority of the loci they identified had only concordant effects and did not assess the effect of the shared loci on individual-level phenotypes. Other studies of genetic overlaps between T2D and CAD have taken a single-disease perspective(23–25,25,27), often decomposing groups of variants into risk pathways using clustering techniques. For example, a recent multi-ancestry analysis of 650 T2D genetic variants with 110 T2D-related traits using bNMF (Bayesian nonnegative matrix factorisation) identified 12 genetic clusters associated with T2D(23) and found 8 of these clusters have different associations with the risk of CAD in individual-level data. As these studies focused on the genetic and biological pathways of T2D, their scope was limited to comprehensively examine the shared biological pathways between both diseases.

In this study, we directly start from a multi-disease perspective, considering both CAD and T2D as main phenotypes and comprehensively identified concordant as well as discordant loci shared between T2D and CAD by using PLACO (pleiotropic analysis under composite null hypothesis)(29) and leveraging power from large multi-ancestry GWAS meta-analysis studies. PLACO is a tool that is designed to identify shared genetic variants between two diseases and have enhanced power to detect additional shared variants that have not reached genome-wide significance in one or either of the individual disease GWAS with maintained type 1 error(29). We then clustered shared variants with intermediate cardiometabolic traits to understand shared pathways and evaluated the effect of these shared groups of variants in UK Biobank (UKB) participants. We find that liver fat and the handling of hepatic lipids partition the shared genetic architecture of T2D and CAD into distinct, mechanistically interpretable risk profiles. Findings from this study offer additional insights into the shared mechanisms underlying T2D and CAD and have implications for precision prevention and for therapies targeting T2D, CAD and metabolic dysfunction-associated steatotic liver disease (MASLD).

## Material & Methods

### Data sources

GWAS summary statistics: For T2D, we used summary statistics from a multi-ancestry GWAS meta-analysis by Mahajan et al. (2022)(30) comprising of 180,834 cases and 1,159,055 controls across 5 major ancestry groups: European, East Asian, South Asian, African and Hispanic participants. We used the effect estimates, standard errors (SEs), and P values from a fixed-effects meta-analysis, which were double-corrected for genomic control. For CAD, we extracted the summary statistics from CARDIoGRAMplusC4D(31), a multi-ancestry GWAS meta-analysis comprising 210,842 cases and 1,167,328 controls from participants of majority European or East Asian ancestry. We used the effect estimates, SEs, and P values from the inverse-variance-weighted meta-analysis and the effect allele frequency (EAF) reported in the summary statistics. In the sensitivity analyses for PLACO and colocalisation, we used GWAS summary statistics from the same studies, restricted to European-ancestry participants for T2D and to those with majority (>97%) European ancestry for CAD (excluding Biobank Japan).

### Quality control (QC) of GWAS summary statistics for analysis using PLACO

Quality control was performed on each set of summary statistics before running PLACO(29) to test for shared variants between T2D and CAD **(Supplementary Figures 1-2)**. We removed INDELs, multi-allelic SNPs (single nucleotide polymorphisms) and SNPs with EAF ≤ 0.01 or ≥ 0.99, as rare variants may violate the asymptotic normality assumption of the maximum likelihood estimator, under which PLACO may fail to control type I error. We merged the two datasets based on chromosome and position and harmonised them to the same effect allele and other allele. We removed SNPs with unresolvable allele mismatches between the datasets and SNPs with squared Z-scores (effect estimate divided by SE)>80 (P≈5×10^-19^) in either dataset, as recommended by the PLACO developers(29) to control the false-positive rate, because these SNPs could disproportionately affect the analysis. This left 8,815,702 variants for the PLACO analysis. To then reduce bias that may arise from sample overlap-induced correlation between the T2D and CAD GWAS, we calculated the Pearson correlation between the Z-scores for the two diseases using genetic variants with ‘no effect’ on either disease (single disease P> 5 × 10^-4^) and decorrelated the Z-scores by using the formula described in the **Supplementary Methods.**

### Characterisation of PLACO-implicated loci

To characterise the shared genetic variants identified by PLACO into genetic loci, variants with PLACO P < 5×10^-8^ were clumped at ±500kb radius according to their linkage disequilibrium (LD), quantified as r^2^, threshold >0.2 (reference panel: 1000G phase 3 All) using FUMA (SNP2GENE function)(32). MHC regions (chr6:28477897-33448354 [genome build 37]) were removed. A locus was defined from independent significant SNPs by merging clumps if they were less than 500kb apart. We represented each locus by the SNP with the lowest P-value. We also redefined genetic loci in the original T2D and CAD GWAS using the same thresholds and method for reporting purposes, so that the locus definitions were consistent with the PLACO results.

As inputs to PLACO, we removed SNPs with large effect sizes (Z-score squared > 80) to control the false-positive rate, as these SNPs could disproportionately affect the analysis. However, to avoid missing any potential shared loci that have variants P < 5×10^-8^ in both the T2D and CAD GWAS but were excluded due to having the large effect sizes, we applied two complementary approaches to try to recover them post-PLACO. First, at the loci-level, we used a Venn diagram and identified any loci that overlapped between the T2D and CAD GWAS but not in PLACO. Second, at the variant-level, we identified whether there were any variants that had P < 5×10^-8^ in both disease GWAS but were not within a PLACO-implicated genetic region. Using these approaches, we recovered 1 locus.

We summarised the direction of effect of SNPs on the risk of T2D and CAD. A locus was defined to have a concordant effect when the effect allele of all PLACO significant SNPs in the region increased or decreased the risk of T2D and CAD at the same time. Discordant effect was defined as all PLACO-significant SNPs in the region increasing the risk of one disease but decreasing the risk of the other. The remaining loci were considered to have mixed effects as they contained variants with both concordant and discordant effects.

We defined a shared locus as previously unreported from two comparisons: 1. We extracted separate lists of variants with P < 5×10⁻⁵ for T2D and CAD from the GWAS Catalog(33) (as of 10^th^ June 2024). We first checked, by distance, whether there were any SNPs with the same rsID or chr:pos in the T2D and CAD variant lists within a ±500 Kb radius of the PLACO index SNP (the SNP with the lowest P value). In the remaining loci, we assessed LD (r^2^>0.2) between all T2D variants and the PLACO index SNP, between all CAD variants and the PLACO index SNP, and between all T2D and CAD variants. A locus was previously unreported if there was no ‘previously associated signal’ in LD with the PLACO index SNP. ‘Previously associated signal’ here refers to a single SNP associated with both CAD and T2D or a CAD and a T2D SNP in LD with each other. 2. We additionally compared our loci with Li et al. (2024)(28) by searching whether any of their shared signals (chr:pos) were inside our genomic regions.

### Colocalisation

To provide further support for sharing between T2D and CAD at PLACO-identified loci, we performed colocalisation analysis. As colocalisation requires a genetic region to be specified, we used the genomic risk locus ranges from clumping in FUMA. For any loci that only had a single fixed position, we added -/+ 500kb from that position. We employed two complementary colocalisation methods: HyPrColoc(34) and prop.coloc(35). HyPrColoc is a Bayesian colocalisation method that estimates the posterior probability that a set of traits share a causal variant within a genetic region. It has the advantage of being robust to sample overlap between GWAS summary statistics. To relax the HyPrColoc’s assumption of a single causal variant in each region, we additionally used GCTA-COJO(36). We first used GCTA-COJO (P value threshold for conditional independent variant = 5×10^-6^) to identify conditionally independent signals for T2D and CAD respectively in each region. We did not set the P value threshold to 5×10^-8^ as there were regions with only genome-wide suggestive significance (P < 5×10^-5^) in the disease GWAS but PLACO significant, thus 5×10^-8^ in GCTA-COJO was too conservative in our case. After GCTA-COJO, we used HyPrColoc (parameters: uniform priors, prior probability=1×10^-4^) to test colocalisation between the conditionally independent signals using the conditioned effect estimates, SEs and P values. Unconditioned (i.e., normal HyPrColoc) was also performed. We defined colocalisation between T2D and CAD in a genetic region with posterior probability >0.75. In addition to testing colocalisation between traits, HyPrColoc also estimates the most likely causal variant shared across the region being tested. We accepted the causal variant when the posterior probability explained by that variant was >0.75.

We additionally employed prop.coloc(35), a frequentist method that tests for proportional colocalisation, a specific type of colocalisation where the associations of causal variants with one trait are proportional to their associations with the other trait. It has the potential to identify colocalisation pattern in regions where HyPrColoc may be limited, such as regions with weaker genetic signals i.e., disease GWAS P value < 5×10^-4^ but are PLACO significant or complex regions that may be missed from the Bayesian framework. For prop.coloc, we used a one-sample setting, given the high degree of sample overlap between our GWAS summary statistics (recommended by the prop.coloc authors). Also recommended by the authors, we set the pruning threshold to 0.4. We defined colocalisation in a region if either the “prop-coloc- cond” or “prop-coloc-full” test passed the colocalisation threshold: p_LM (Lagrange Multiplier test) < 0.05 and p_coloc > 0.05.

For GCTA-COJO and prop. coloc, we used genotype data from unrelated UKB participants of European ancestry as a reference panel to calculate the LD matrix.

### Clustering analysis

To understand how shared variants might mediate risk of both T2D and CAD, we clustered these variants based on their cardiometabolic trait associations using Bayesian nonnegative factorisation (bNMF), adapting a previously developed QC and analysis pipeline(23,25,26). Variant selection and data pre-processing steps for clustering are detailed in **Supplementary Figures 3 and 4.** To select variants for the clustering, we extracted all PLACO-significant variants, variants not tested in PLACO, and 24 potentially causal variants identified in colocalisation. We removed SNPs in the MHC region and applied LD clumping (kb=500, r^2^=0.05, P=5×10^-8^, MAF=0.01) and LD pruning (50, 5, 0.05, MAF=0.01) using UKB European participants as the LD reference panel. These steps produced a final list of 187 independent variants (**Supplementary Table 1**) for clustering.

For traits included in the clustering, we first prioritised 55 cardiometabolic risk factors that were included in previous T2D loci clustering papers (23,24). To expand the traits set, we used a PheWAS approach to look up variant-trait associations in the OpenTargets database(37), and included any traits that have P < 5×10^-5^ with at least two of the 187 variants. When choosing the GWAS summary statistics for each trait, we prioritised multi-ancestry GWASs; if a multi-ancestry GWAS was unavailable, we used the largest European GWAS. Traits were removed if their median GWAS sample size was <5,000 or if their minimum P value in the final variant set was not Bonferroni significant (P_min_ > 0.05/187 variants) or if they were highly correlated with other traits (Pearson correlation coefficient > 0.8). After exclusion, the final set comprised 77 traits (**Supplementary Table 2**). We filled in any missing variant-trait Z-scores with a cover proxy (LD r^2^>0.5) or the trait’s median variant Z-score among the 187 variants if an LD proxy was not available. We standardised based on the GWAS sample size and scaled Z-scores, which had been aligned to the T2D risk-increasing alleles.

We ran bNMF using iteration=10000, tolerance=1×10^-6^, K0=2, K=10(23). Top-weighted variants defined the clusters, and we employed the method used in Kim et al. (2024)(26) to determine an optimal weight cut-off that maximised the signal-to-noise ratio. The cut-off was 1.2181 in this analysis.

We compared our clusters with Smith et al. (2024)(23) and Suzuki et al. (2024)(24). For each of our clusters, we calculated the percentage overlap of variants and traits with each cluster identified in the two studies. For variants, we divided the number of top-weighted variants in our cluster that are the same or in LD (r^2^ > 0.5) with variants in either previous study by the total number of top-weighted variants in our cluster. We applied the same method for comparing traits. We gave our clusters the same names as Smith et al. (2024)(23) for reporting and comparison purposes when the key variants/traits aligned.

### Metabolic traits analysis

Given the metabolic aetiology of T2D and CAD, we conducted a metabolic traits analysis to gain a deeper insight into the cluster’s associations with metabolic traits. We used GWAS summary statistics for 140 metabolic traits(38) from the Nightingale Health NMR panel and 185/187 variants from the clustering. Two variants (GRCh38: 12:109275606_G_A, 12:109538471_G_A) were excluded because they were missing in all 140 metabolite GWAS. For the remaining 185 variants, we filled any missing values with LD proxies (same selection as in the clustering method) or, if not available, with the metabolite’s median variant Z-score among the 185 variants. We standardised based on GWAS sample size and scaled Z-scores, which had been aligned to the T2D risk-increasing alleles. We coded a binary variable for each cluster, with the top-weighted variants coded 1 and 0 otherwise and assessed the combined effect of the top-weighted variants in each cluster on the metabolic traits adjusted for other clusters by using linear regression:

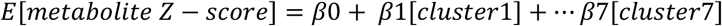

We adjusted for multiple testing by using 0.05/28 (reported number of metabolite PCs (principal components) that explained 95% of the variation in the metabolic traits(38)).

### Individual-level analysis in the UK Biobank cohort

To replicate findings from the clustering and metabolic trait analysis in individual-level data, for each UKB participant, we calculated a genetic risk score (GRS) for each of the 7 clusters and assessed the association between the cluster GRS and 17 clinical biomarkers, T2D, CAD as well as 1,254 other binary outcomes. Sample exclusion and analysis design are shown in **Supplementary Figure 5**. The analysis included 378,772 individuals after excluding participants that have withdrawn (n=3), without genetic data (n=15,204), non-European ancestry (n=78,371), sex mismatch (n=308), related participants (KING kingship coefficient cutoff=0.0884; n=29,704) and missing from the Phecode dataset (n=4). Baseline characteristics are shown in **Supplementary Table 3**.

All clinical biomarker data were taken from baseline; prevalent and incident disease status was defined using a combination of self-reported health history, medication status, and linked diagnosis and procedure codes using established definitions of CAD(39) (N controls=340,239, N cases=38,533) and T2D(40) (N controls=343,950, N cases=32,869). Cases for the 1,254 other binary outcomes were defined as Phecodes (v1.2) using condition-specific ICD-9/10 codes in linked HES, cancer and death registry datasets(41). We removed binary outcomes with <100 cases and included both prevalent and incident cases in the analysis.

We calculated the cluster GRS by summing the product of the number of T2D-increasing alleles and their cluster weights across the top-weighted variants that defined the cluster. We used the cluster loading weights instead of effect estimates from the T2D GWAS to avoid overfitting. The cluster GRS was then standardised to a mean of 0 and SD of 1. We assessed associations between cluster GRS and 17 clinical biomarkers (key traits in the clustering analysis) using linear regression. We adjusted for antihypertensive use by adding 15 and 10 mmHg to participants’ systolic and diastolic blood pressure (SBP, DBP) levels, respectively (42). For participants on lipid-lowering medication, we multiplied their total cholesterol levels by 1.25, LDL (low-density lipoprotein) by 1.43, and triglycerides by 1.18 (23,43). To reduce the influence of outliers, all variables were winsorised at 5 SDs from the mean (27). To meet the homoskedasticity assumption for linear regression, we natural log transformed BMI (body mass index), HDL (high-density lipoprotein), Trig (triglycerides), CRP (C-reactive protein), ALP (alkaline phosphatase), ALT (alanine aminotransferase), GGT (gamma-glutamyl transferase), AST (aspartate aminotransferase), liver fat PDFF (proton density fat fraction) and SHBG (sex hormone-binding globulin). Associations with T2D, CAD and the 1,254 binary outcomes were assessed using logistic regression. Analyses were adjusted for age, sex (sex not included for binary outcomes that affect only one sex), and the first 5 genetic PCs. Multiple testing was corrected using the Bonferroni method, i.e., for each cluster, we used 0.05 divided by 17 (the number of biomarkers) and 0.05 divided by 1,254 (the number of binary disease outcomes tested).

### Two-sample Mendelian randomisation

To assess whether the relationship between liver fat (exposure) and T2D, and liver fat and CAD may be causal, we performed two-sample MR(44–46). For liver fat, we took 25 independent variants (**Supplementary Table 4**) identified in Björnson et al. (2024) (UKB, EUR)(47) and used the effect estimates from an external liver fat GWAS (Ahmed et al. (2025)(48), as full GWAS summary statistics was not available in Björnson et al. (2024). The external liver fat GWAS was also conducted in UKB and using the same phenotype. To avoid sample overlap with the outcome GWAS, for CAD, we used the GWAS summary statistics from FinnGen Release 13 (I9_CHD)(49) and for T2D, Mahajan et al. (2018) (50) excluding UKB. To further understand whether ApoB-containing lipoproteins may be mediating the relationship between liver fat and CAD as well as T2D, we conducted partitioned two-sample MR where the 25 liver fat variants were partitioned into 3 groups derived by Björnson et al. (2024): lipoprotein-increasing (NSNP=3), lipoprotein-neutral/mixed (NSNP=12), and lipoprotein-decreasing (NSNP=10) based on the variants’ effect on ApoB, LDL, triglycerides and TRL-C (Triglyceride-Rich Lipoprotein Cholesterol). We used the effect estimates from Ahmed et al. (2025)(48) to replicate the partitioned MR results for CAD in Björnson et al. (2024) and additionally extend them to T2D.

In all two-sample MR analyses, alleles were aligned to liver fat-increasing alleles, and effect estimates were harmonised between the exposure and outcome datasets. We used the inverse-variance weighted (IVW) method(51,52) as the main MR method in our analysis when there was no statistical evidence of horizontal pleiotropy, assessed using the MR-Egger intercept (pleiotropy test)(46). If the pleiotropy test was significant (P < 0.05), we used the results from MR-Egger(46), which produces unbiased estimates when instruments have horizontal pleiotropy. We also performed 2 other pleiotropy-roust methods: weighted median(53) and the weighted mode(54) as additional sensitivity analyses to MR-Egger. To assess whether there were any single instrument driving the results and the robustness of the findings overall, we further conducted leave-one-out and single-SNP analyses.

### Software and packages

Analyses were conducted using R version 4.3.1 and the following packages: PLACO (version 0.1.1), GenomicRanges (version 1.54.1), GCTA (version 1.94.3), HyPrColoc (version 0.0.2), prop.coloc (version 1.1.0), ieugwasr (version 1.0.1), adapted codes from Kim et al 2023(26) for the bnmf clustering pipeline (release Nov 25, 2024), ukbtools (version 0.11.3), speedglm (version 0.3.5), pheatmap (version 1.0.12), ggplot2 (version 3.5.0), TwoSampleMR (version 0.6.8, R version 4.4.2), PLINK (v1.90b7) and PLINK2 (v2.00a6LM).

## Results

### Identification of shared genetic loci between T2D and CAD

Recent large-scale multi-ancestry GWAS meta-analyses discovered 260 and 284 independent loci for CAD(31) and T2D(30) respectively **(Supplementary Table 5)**. To identify shared loci, we applied PLACO(29) to these data, identifying a total of 149 shared genetic loci **(Figure 1, Supplementary Table 6, Supplementary Figure 6)**, of which 42 were previously unreported shared loci. Among 149 shared loci, 40 were significant (P<5×10^-8^) in both the T2D and CAD GWAS, 61 were only significant in the T2D GWAS, 38 were only significant in the CAD GWAS and 10 were not significant in either disease GWAS. Sensitivity analysis using GWAS summary statistics restricted to European ancestry participants identified 101 shared loci between CAD and T2D, of which 90 (89.1%) were also identified in the main analysis **(Supplementary Table 7)**. Among the 149 loci in the main analysis, 73 loci had concordant effects on the risk of T2D and CAD, 57 had discordant effects, and 19 had mixed effects **(Supplementary Table 6)**.

**Figure 1.**
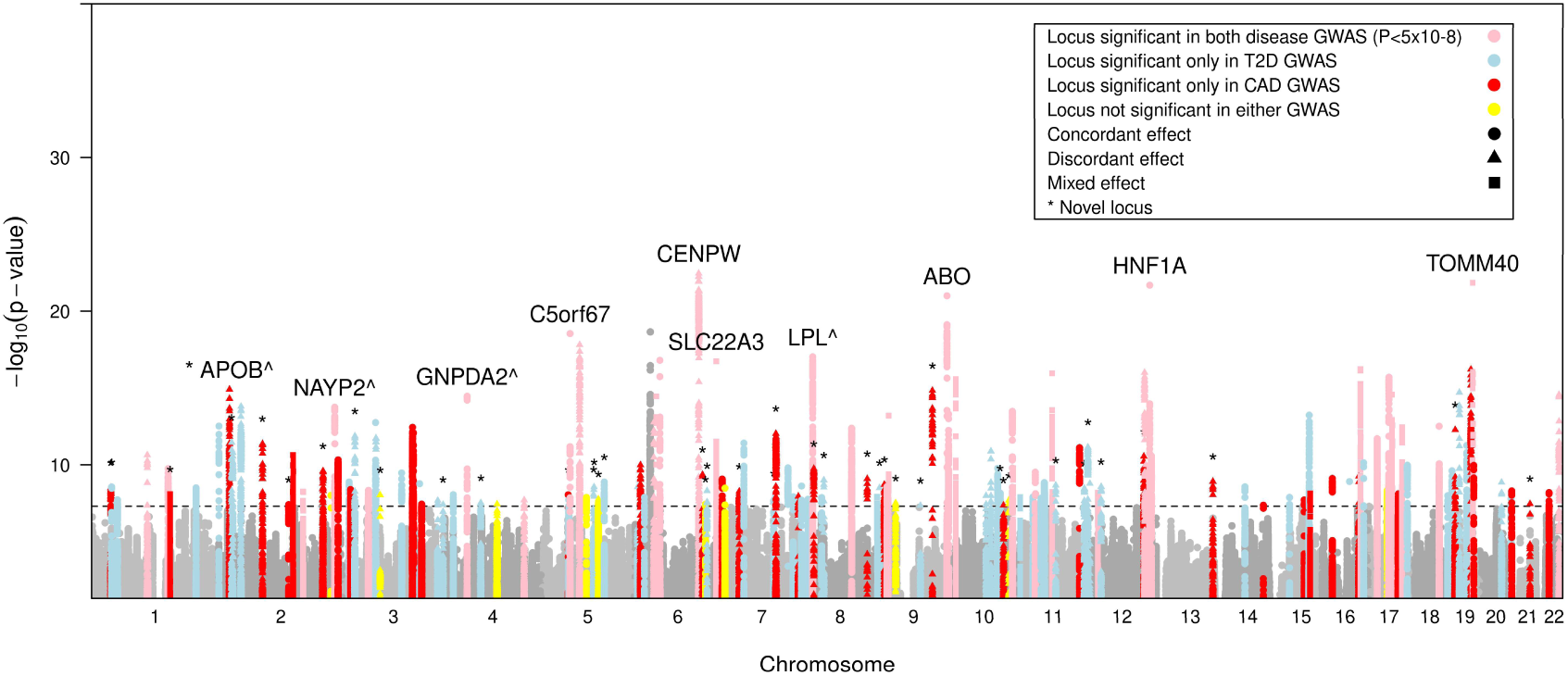
Shared genetic loci between T2D and CAD. Manhattan plot illustrating the significant genetic regions for shared genetic loci between type 2 diabetes and coronary artery disease identified by PLACO. The dashed horizontal line marks the genome-wide significance threshold (P=5×10^-8^ or 7.30 in the -log10 scale). The nearest protein-coding gene is also labelled (*^*).

To provide orthogonal evidence that PLACO-identified loci were shared between T2D and CAD, we conducted colocalisation analyses using two methods with differing assumptions (COJO-HyPrColoc(34,36) and prop-coloc(35). Using multi-ancestry GWAS data and a UKB European LD reference panel, we identified 98 genetic loci with additional support from at least one colocalisation method **(Figure 2, Supplementary Table 8)**. We conducted sensitivity analyses to compare colocalisation between using summary statistics derived from multi-ancestry cohorts and European cohorts. Results showed a high degree of consistency across both analyses, suggesting that the likelihood of false-positive findings due to differences in LD in our main colocalisation analysis was low (**Supplementary Results, Supplementary Table 8, Supplementary Table 9**).

**Figure 2.**
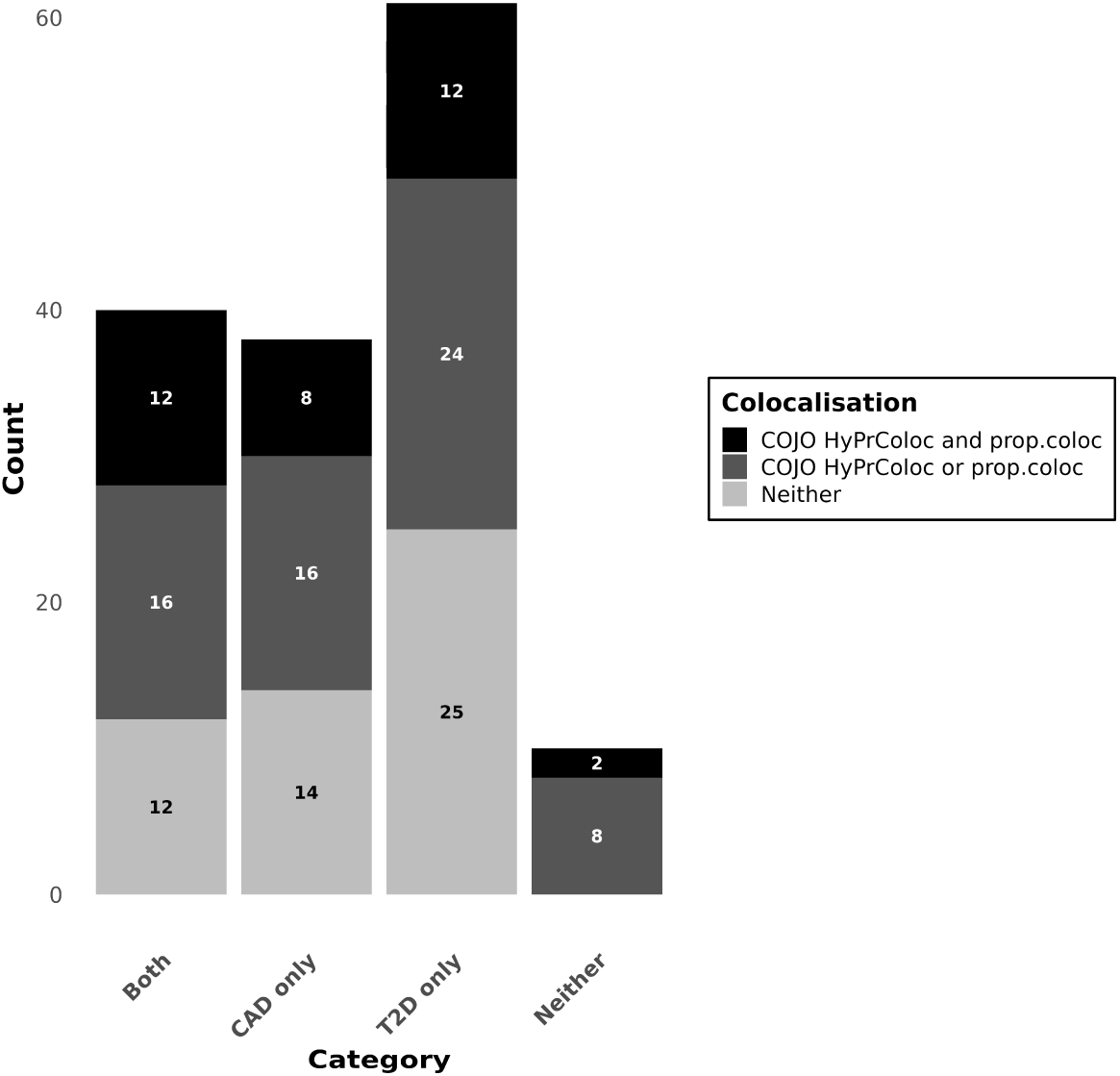
Number of shared genetic loci between T2D and CAD with colocalisation evidence. Bar chart summarising the number of shared genetic loci (149 in total) with colocalisation support from COJO HyPrColoc, prop.coloc, both or neither. The category indicates whether a genetic locus was originally significant in both the CAD and T2D GWAS, in either GWAS alone, or in neither.

### Clustering analysis of shared variants to identify common biological pathways between T2D and CAD

To understand how shared variants might mediate risk of both T2D and CAD, we clustered genetic variants based on their associations with intermediate phenotypes, including known risk factors. We adapted the bNMF method, which has been previously applied to T2D(23,24), to a comorbidity context using shared variants for T2D and CAD. After clustering 187 shared variants (aligned to the T2D-increasing allele) with 77 intermediate traits, we identified 7 clusters (including 68 and 46 unique top-weighted variants and traits, respectively) shared between T2D and CAD **(Figure 3, Supplementary Table 10)**. Clustering the same variants and traits with alleles aligned to the CAD-increasing allele produced nearly identical results **(Supplementary Figure 7, Supplementary Table 11)**. We found 4 clusters (*ALP negative, Lipodystrophy 1, Beta Cell 2,* and *Obesity*) defined primarily by concordant variants, and 3 clusters (*Cholesterol, Liver fat (Retention),* and *Liver lipid*) defined primarily by discordant variants (**Supplementary Table 10**). The naming of our shared T2D-CAD clusters reflected the partial overlap with previously reported T2D clusters (23,24) (**Supplementary Table 12**).

**Figure 3.**
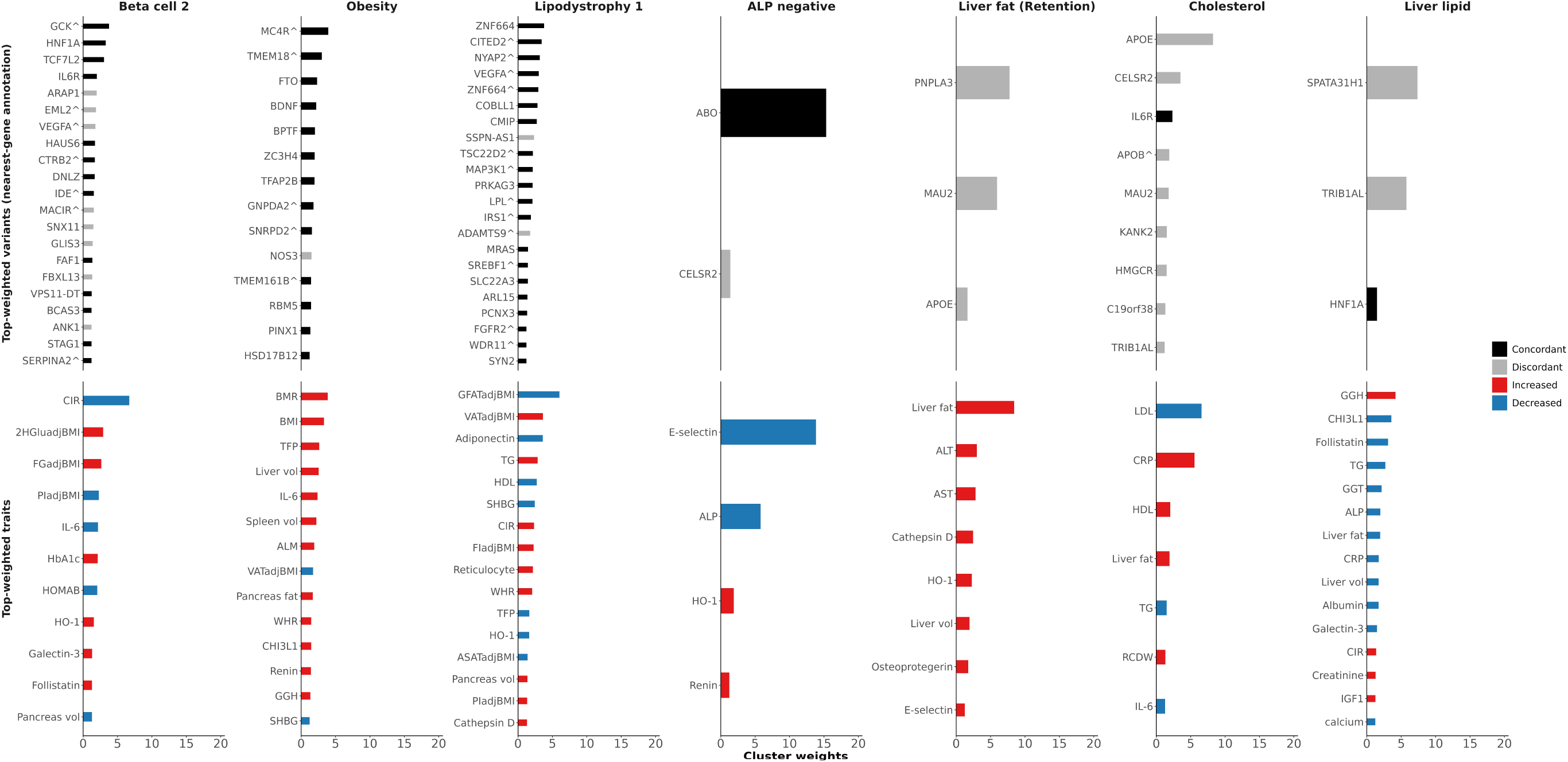
Clustering analysis identifies 7 shared clusters of genetic variants between T2D and CAD. Figure illustrates the defining variants and traits for each genetic cluster shared between T2D and CAD. Alleles aligned to T2D-increasing alleles. ^nearest protein-coding gene. 2HGluadjBMI, two-hour glucose adjusted for BMI; FGadjBMI, fasting glucose adjusted for BMI; HO-1, heme-oxygenase 1; BMR, basal metabolic rate; TFP, trunk fat percentage; ALM, Appendicular lean mass; GGH, Gamma glutamyl hydrolase; VATadjBMI, Visceral adipose tissue volume adjusted for BMI and height; TG, triglycerides; FIadjBMI, fasting insulin adjusted for BMI; PIadjBMI, proinsulin adjusted for BMI; GFATadjBMI, Gluteofemoral adipose tissue volume adjusted for BMI and height; RCDW, Red cell distribution width; creatinine, serum creatinine; albumin, serum albumin

To increase confidence in our clustering results, we assessed the fraction of variants in each cluster that were in regions which colocalised between T2D and CAD. For the *ALP negative* and *Liver fat (Retention)* clusters, all variants were in colocalising regions. The variants that defined the *Liver fat (Retention)* cluster were also the shared causal variants prioritised by colocalisation. Other clusters also had high proportions of variants in each cluster that colocalised (*Beta cell 2* – 13/21 (61.9%), *Cholesterol* – 6/9 (66.7%), *Lipodystrophy 1* – 17/22 (77.3%), *Obesity* – 10/14 (71.4%)), suggesting that the clustering captured genuine overlapping pathways between T2D and CAD. Only the *Liver lipid* cluster had a relatively lower colocalisation proportion (33.3%, **Supplementary Table 13**).

We identified three clusters associated with liver fat with discordant effects on T2D and CAD. The *Liver fat (Retention)* cluster included three variants at genes *PNPLA3*, *MAU2* and *APOE* involved in hepatic lipid regulation and cholesterol/fat accumulation(55) where the T2D risk allele was associated with increases in liver fat, liver volume, and liver enzymes **(Figure 3)**. The top-weighted variant, rs738408, is in perfect LD (r^2^=1) with a well-characterised missense variant: rs738409, in *PNPLA3* (I148M), which is associated with risk of T2D, non-alcoholic fatty liver disease (NAFLD), but reduced risk of CAD, potentially due to impaired intrahepatic triglyceride breakdown and reduced VLDL secretion(55–58). rs73001065 (*MAU2*) is in high LD (r^2^=0.88) with a well-known missense variant, rs58542926, in *TM6SF2* which affects hepatic triglyceride content and is associated with T2D, NAFLD and CAD in the same directions as rs738409(55,59–61). rs429358 is a missense variant in the *APOE* gene, which plays a key role in cholesterol and triglyceride metabolism in the brain and liver. The T allele had been found to be associated with increased risk of T2D and liver fat, and a decreased risk of coronary artery disease(57).

The *Cholesterol cluster* contained the same variants at the *MAU2* and *APOE* genes, along with 4 additional colocalised variants. rs12740374 is a 3′-UTR variant in the *CELSR2* gene, but it is known to regulate *SORT1* expression in the liver. The other three colocalised variants are near genes *HMGCR*, *KANK2* and *C19orf38* (a poorly characterised gene). *CELSR2- SORT1*, *HMGCR* and *KANK2* are involved in the regulation of lipid metabolism and cholesterol synthesis across the liver and other tissues, but do not as strongly affect hepatic lipid transport and fat storage compared to rs738408/rs738409 in *PNPLA3.* Unlike the *Liver fat* (Retention) cluster, the *Cholesterol* cluster did not contain the PNPLA3 variant, nor were liver enzymes or liver volume top-weighted traits in this cluster **(Figure 3)**.

The *Liver lipid* cluster was associated with lower levels of liver fat, liver volume, and liver enzymes (ALP and GGT), in directions opposite to the two previous clusters **(Figure 3)**. The top-weighted variant was rs4665988, an intronic variant in *SPATA31H1* that is in moderate LD (r^2^=0.41) with rs1260326, a missense variant in *GCKR* that affects the ability of *GCKR* to inhibit glucokinase and thereby glucose and hepatic lipid/fat storage(62). rs1260326 has consistently been reported to have an opposite direction of effect between T2D and liver fat(55,62–64) and the T2D risk allele was weakly associated with decreased risk of CAD(57). The second top-weighted variant rs2954022 is an intron variant in *TRIB1AL*, a long non-coding RNA located adjacent to *TRIB1*. *TRIB1* is a well-characterised protein-coding gene that plays a role in hepatic lipids metabolism and atherosclerosis(65). rs2954022 is in near perfect LD (r^2^=0.97) with rs6982636 (intron variant overlapping with *TRIB1AL*), where the A allele was associated with lower liver fat and risk of CAD (31,48); however, no association between rs6982636 and the risk of T2D have been reported to date.

Given the metabolic aetiology of both diseases, we additionally evaluated the effect of each genetic cluster on 140 circulating metabolic traits from the Nightingale Health NMR panel using GWAS summary statistics(38). In the *Liver fat (Retention)* cluster, we found a lipid profile characterised by reduced LDL and VLDL (very low-density lipoprotein) traits and elevated HDL traits, representing a classic cardioprotective pattern and consistent with the direction of CAD association from the clustering analysis **(Figure 4, Supplementary Table 14)**. The *Cholesterol* cluster showed a similar pattern but with a larger decrease in IDL (intermediate-density lipoprotein) and LDL traits and was additionally associated with decreases in some HDL traits, sphingomyelins, phosphatidylcholine, total cholines, and phosphoglycerides **(Figure 4, Supplementary Table 14)**. The *Liver lipid* cluster was associated with decreases in VLDL (but not very small VLDL) traits and increases in large HDL traits **(Figure 4, Supplementary Table 14)** but no net effect on LDL.

**Figure 4.**
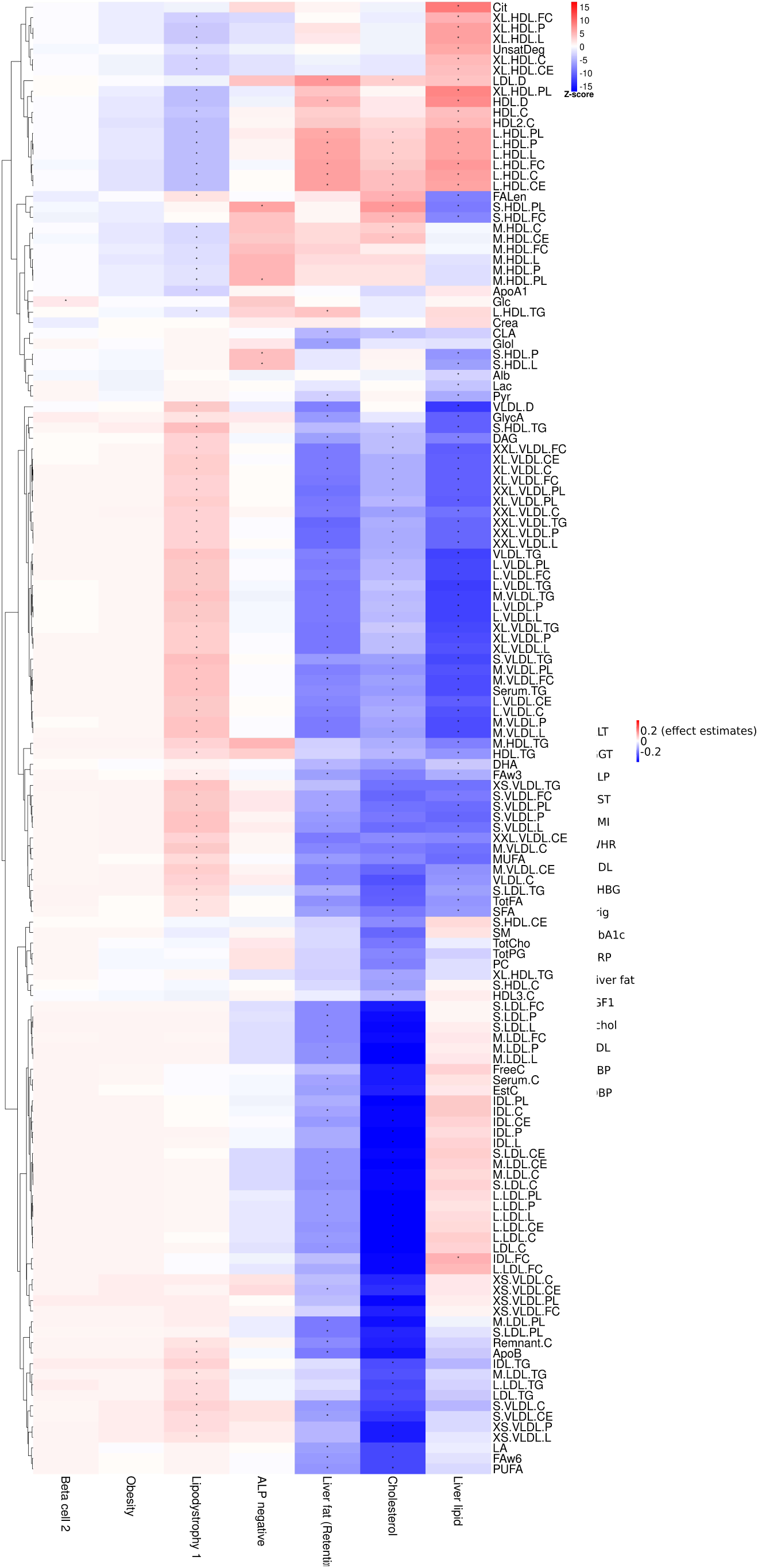
Heatmap of associations of the combined effect of top-weighted variants in shared genetic clusters between T2D and CAD with metabolic traits. Effect estimates and P values were derived from linear regression by regressing Z-scores (beta/SE from GWAS summary statistics) of metabolites over cluster x (coded as a binary variable: 1 for top-weighted variant and 0 otherwise), adjusted for other clusters. Red indicates that the overall effect of alleles of the variants in the cluster has an increasing effect on the metabolic trait, and blue indicates a decreasing effect. Asterisks denote statistically significant results that met the multiple testing threshold (p<0.05, corrected for the number of independent metabolite PCs).

The other four clusters, which were defined primarily by concordant variants, also exhibited distinct patterns of association with metabolic traits **(Figure 4, Supplementary Table 14)**. The *ALP negative* cluster showed an increase in phospholipids in medium and small HDL particles, and in the concentration and total lipids in small HDL. *The Beta Cell 2* cluster showed no notable increases in metabolites other than glucose, consistent with the functional role of pancreatic tissue and beta cells in glucose metabolism. The *Lipodystrophy 1 cluster* showed a metabolic profile characteristic of elevated CAD risk characterised by elevated LDL and VLDL and reduced HDL. In the *Obesity* cluster, there was no net changes in metabolic traits.

### Associations of cluster GRS with biomarkers and disease outcomes in the UKB

To validate patterns observed in the clustering analysis in individual-level data, we assessed the association between cluster GRS and T2D, CAD, and a range of clinical biomarkers and binary outcomes in the UK Biobank. We found that all cluster-specific GRS were significantly associated with the risk of T2D and CAD except the *ALP negative* cluster and were consistent with the clustering results **(Table 1)**. For example, the *Liver fat (Retention)* and the *Cholesterol* cluster were significantly associated with increased T2D risk (*Liver fat (Retention)* OR: 1.03 [1.02 – 1.04], P=2.42×10^-7^, *Cholesterol* OR: 1.06 [1.04 – 1.07], P=3.06×10^-19^) and decreased CAD risk (*Liver fat (Retention)* OR: 0.98 [0.97 – 0.99], P=5.67×10^-5^, *Cholesterol* OR: 0.93 [0.92 – 0.94], P=2.46×10^-41^. The *ALP negative* cluster was not significantly associated with increased risk of CAD (OR 1.01 [1.00-1.02], P=0.13) **(Table 1)**.

**Table 1.** Effect of cluster GRS on the risk of T2D and CAD in the UK Biobank. Effect estimates derived from a logistic regression model adjusted for age, sex and genetic principal components 1-5. Multiple testing correction was not applied. T2D and CAD included both incident and prevalent cases.

| Cluster GRS | Majority cluster-defining variants | T2D |  | CAD |  |
| --- | --- | --- | --- | --- | --- |
|  |  | OR | P value | OR | P value |
| ALP negative | Concordant | 1.03 (1.02-1.04) | $4.38 \times 10^{-8}$ | 1.01 (1.00-1.02) | 0.13 |
| Beta cell 2 | Concordant | 1.16 (1.14-1.17) | $5.74 \times 10^{-137}$ | 1.04 (1.03-1.05) | $6.98 \times 10^{-14}$ |
| Lipodystrophy 1 | Concordant | 1.13 (1.12-1.14) | $3.16 \times 10^{-97}$ | 1.06 (1.05-1.07) | $6.84 \times 10^{-28}$ |
| Obesity | Concordant | 1.09 (1.08-1.11) | $7.27 \times 10^{-53}$ | 1.04 (1.03-1.05) | $5.11 \times 10^{-13}$ |
| Cholesterol | Discordant | 1.06 (1.04-1.07) | $3.06 \times 10^{-19}$ | 0.93 (0.92-0.94) | $2.46 \times 10^{-41}$ |
| Liver Fat (Retention) | Discordant | 1.03 (1.02-1.04) | $2.42 \times 10^{-7}$ | 0.98 (0.97-0.99) | $5.67 \times 10^{-5}$ |
| Liver lipid | Discordant | 1.02 (1.01-1.04) | $6.31 \times 10^{-5}$ | 0.97 (0.96-0.99) | $1.70 \times 10^{-6}$ |

In the clinical biomarker analysis, the pattern of associations was also consistent with the clustering results **(Figure 5A)**. In addition to the *Liver fat (Retention), Cholesterol* and *Liver lipid* cluster having associations with liver fat, the *ALP negative* and *Lipodystrophy 1* cluster GRS also had (weaker) associations with liver fat. All cluster-specific GRS, except the *Liver lipid* cluster, were associated with blood pressure, despite a lack of SBP and DBP enrichments among the clustering-analysis traits **(Figure 5A, Supplementary Table 15)**. The direction of the effect on blood pressure also differed between clusters. For example, *Cholesterol, Lipodystrophy 1, Liver fat (Retention),* and the *Obesity* cluster were associated with increased DBP, while the *ALP negative* cluster was associated with decreased DBP. The *Liver fat (Retention)* cluster showed a greater increase in liver fat. In contrast, the *Cholesterol* cluster showed a much greater decrease in triglycerides, LDL, and total cholesterol compared to the *Liver fat (Retention)* cluster. The *Cholesterol* cluster was also linked to slightly increased BMI and WHR (waist-hip-ratio), while the *Liver fat (Retention)* cluster was not associated with BMI. In contrast to these two clusters, the *Liver lipid* cluster was associated with decreased liver fat but increased IGF-1, as expected and consistent with the clustering results **(Figure 5A, Supplementary Table 15)**.

**Figure 5.**
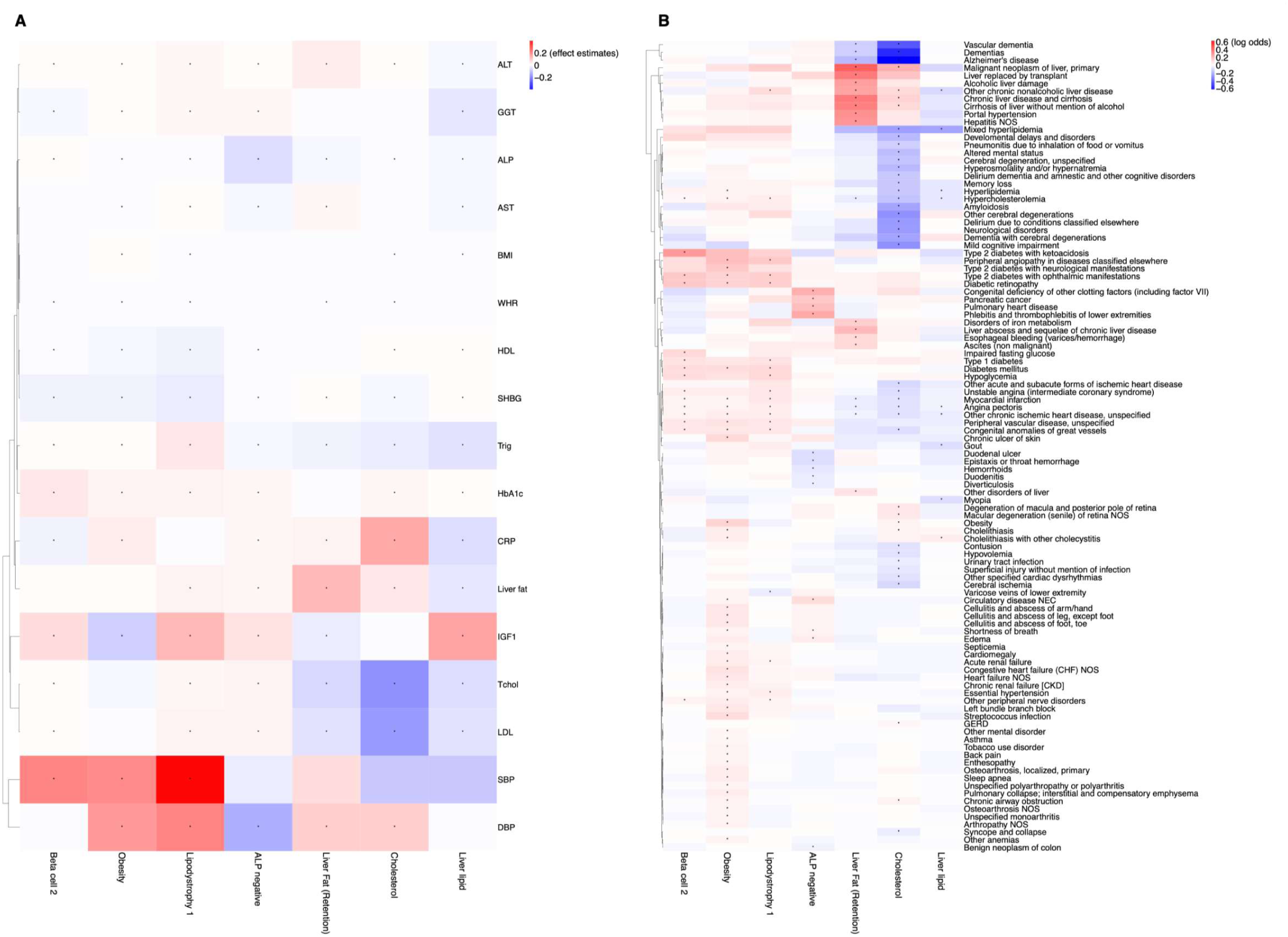
Effect of cluster-specific GRS on clinical biomarkers and binary outcomes in the UK Biobank. Effect estimates were derived using linear regression (for continuous biomarkers) and logistic regression (for binary outcomes), adjusted for age, sex, and the first 5 genetic principal components. Multiple testing was corrected using the Bonferroni method. (**A**) Effect of cluster GRS on biomarkers. (**B**) Effect of cluster GRS on binary outcomes (figure visualisation restricted to outcomes that passed the Bonferroni multiple testing threshold). GRS, genetic risk score; IGF1, insulin growth factor-1; SHBG, Sex Hormone–Binding Globulin; Tchol, total cholesterol; WHR, waist-hip-ratio; DBP, diastolic blood pressure; SBP, systolic blood pressure.

Clusters also showed distinct patterns of association with binary outcomes **(Figure 5B, Supplementary Table 16)**. As expected, variants in the *Beta Cell* cluster were mainly associated with T2D, T2D complications and heart diseases, supporting the central role of beta cell dysfunction in the pathogenesis of T2D. The *Lipodystrophy 1* cluster also had similar associations with T2D and T2D complications, but was less diabetes dominated and had associations with renal failure and other types of heart and vessel diseases. As expected, the *Obesity* cluster was associated with obesity diagnoses and a much wider range of diseases e.g., from T2D, CAD to cellulitis, renal failure, to asthma, sleep apnoea and osteoporosis, compared with other clusters. As observed in the clustering analysis, the *Liver fat (Retention)* and *Cholesterol* clusters were associated with an increased risk of liver diseases. The *Cholesterol* cluster, in comparison to the *Liver fat (Retention)* cluster, was additionally associated with obesity and a broader range of diseases (e.g. cerebrovascular conditions). The *Liver lipid* cluster, which was associated with decreased liver fat, was also associated with a concomitant decrease in the risk of other chronic non-alcoholic liver disease **(Figure 5B, Supplementary Table 16)**. These results in the UKB validated the patterns seen in the clustering.

### Two-sample MR suggests liver fat causally contributes to the discordant genetic risk between T2D and CAD

To investigate whether the relationships between liver fat and the discordant risk between T2D and CAD may be causal, we conducted a standard two-sample MR analysis. We extracted 25 variants (**Supplementary Table 4**) associated with liver fat (exposure) identified in Björnson et al. (2024)(47) and extracted the effect estimates for these 25 variants from an external study(48), as full GWAS summary statistics was unavailable in Björnson et al. (2024)(47). We observed that genetically increased liver fat was associated with increased risk of T2D (OR=1.309 [1.162–1.474], P=9.52×10^-6^) and decreased risk of CAD (OR=0.893 [0.827–0.964], P=3.77×10^-3^) **(Table 2, Supplementary Table 17-19, Supplementary Figure 8-9),** consistent with findings from our *Liver fat (Retention)* and *Cholesterol* clusters.

**Table 2.** Two-sample MR results between liver fat and CAD and T2D. The table illustrates the causal effects (Odds Ratios and 95% CIs) of liver fat (exposure) on CAD (outcome 1) and T2D (outcome 2), using 25 liver fat variants and variants stratified by their effects on circulating ApoB-containing lipoproteins. We extracted the 25 liver fat variants identified in Björnson et al. (2024) (UKB, EUR)(47) and used the effect estimates from an external liver fat GWAS (Ahmed et al. (2025)(48)), as full GWAS summary statistics was not available in Björnson et al. (2024)(47). Variants stratification by their effects on ApoB-containing lipoproteins also followed Björnson et al. (2024)(47). GWAS summary statistics for CAD and T2D were extracted from FinnGen release 13(49) and Mahajan et al. 2018(50) without UK Biobank respectively. N_SNP_, number of SNPs; IVW, inverse variance weighted.

| Exposure (N<sub>SNP</sub>) | Effect on circulating ApoB-containing lipoproteins | Causal Effect (IVW method) |  |
| --- | --- | --- | --- |
|  |  | CAD | T2D |
| Standard two-sample MR using all variants associated with liver fat |  |  |  |
| Liver fat (25) | - | 0.893 (0.827–0.964) | 1.309 (1.162–1.474) |
| Partitioned MR using liver fat variant clusters defined by their effects on ApoB-containing lipoproteins |  |  |  |
| Liver fat (3) | Increasing | 1.592 (1.066–2.377) | 0.657 (0.262–1.645) |
| Liver fat (12) | Neutral/Mixed | 0.902 (0.846–0.963) | 1.271 (1.153-1.401) |
| Liver fat (10) | Decreasing | 0.817 (0.754–0.885) | 1.527 (1.337-1.744) |

In Björnson et al. (2024)(47), the authors found that the partitioned groups of liver fat variants had opposing effects on the risk of CAD, suggesting that ApoB-containing lipoproteins were driving the effect on CAD, rather than from liver fat directly. To further examine the relationship between liver fat, CAD and T2D, we replicated the partitioned MR results for CAD in Björnson et al. (2024) using effect estimates from the external study(48) and then extended them to T2D. We observed an increase in genetically determined liver fat that is associated with a decrease in ApoB-lipoproteins (this group included variants in our *Liver fat (Retention)* and *Cholesterol* cluster), increased the risk of T2D (OR=1.57 [1.337-1.744], P=4.03×10^-10^) and decreased the risk of CAD (OR=0.817 [0.754-0.885], P=7.94×10^-7^) **(Table 2, Supplementary Table 17-19, Supplementary Figure 10**-17), consistent with our clustering, metabolic traits and UKB results. Increase in genetically determined liver fat that is associated with an increase in ApoB-lipoproteins increased the risk of CAD but was not statistically significant for T2D (**Table 2, Supplementary Tables 17-19, Supplementary Figures 10-17)**. To note, this group of variants only included one variant (rs6982636 and rs2954022 in near perfect LD) from the *Liver lipid* cluster, thus they are related but not identical groupings and this part of the partitioned MR results should not be conflated with the *Liver lipid* cluster. These MR results indicate that while liver fat largely sets the direction of T2D, the direction of CAD is set by how the liver handles the lipids (hepatic lipid retention or impaired lipid export), rather than from liver fat directly.

## Discussion

We identified 149 genetic loci shared between T2D and CAD. Among these, 42 were previously unreported and primarily comprised discordant effects between T2D and CAD. By clustering genetic variants across these 149 loci with cardiometabolic traits, we identified 2 clusters (*Liver fat (Retention)* and *Cholesterol*) associated with increases in liver fat and T2D, but decreased risk of CAD, which were validated in UKB participants. Analysis using metabolic traits showed that the *Liver fat (Retention)* and the *Cholesterol* clusters were associated with decreased VLDL and LDL and increases in HDL, consistent with clustering results indicating decreased risk of CAD. Two-sample MR showed that increase in liver fat was directly associated with T2D risk while the effect on CAD depended on the handling of ApoB-containing lipoproteins by the liver, rather than from liver fat directly.

We replicated many previously reported shared genetic loci between T2D and CAD. In contrast to previous reports(28,66) where most of the shared loci showed directionally concordant effects between T2D and CAD, the majority of the previously unreported loci we identified were discordant. Discordant loci provide further insights into the complex shared aetiology underlying T2D and CAD, revealing biological trade-offs that may inform precision medicine, in which managing one condition might confer risk on another. Discordant loci may have been missed in previous studies due to differences in the GWAS datasets used and the statistical methods applied, which may have preferentially detected concordant loci. Our study using PLACO(29) directly tests pleiotropy at the variant level, leading to an equal chance of detecting concordant and discordant signals. By applying PLACO to large-scale multi-ancestry GWAS data, we enhanced our power to identify additional shared loci between T2D and CAD beyond those previously reported.

We clustered shared variants between CAD and T2D using bNMF to elucidate their common biological pathways further. Our approach differs from previous studies that clustered variants associated with a single disease (23,24,67). We identified 7 genetic clusters, of which 2 (*Liver fat (Retention)* and *Cholesterol*) were defined by increased liver fat and increased T2D risk but decreased CAD risk. The *Liver fat (Retention)* cluster included variants in the *PNPLA3*, *MAU2* and *APOE* genes that are known to be strongly associated with both our focal diseases and with liver fat and steatosis(23,24,31,55,57–59,61). We provide additional support for these associations by showing that CAD and T2D colocalise at these loci and that the variants in this cluster were the likely shared causal variants. Our *Liver fat (Retention)* cluster had a full overlap with the *Liver and lipid Metabolism* cluster in Suzuki et al. (2024)(24); however, the previous study did not specifically explore the cluster’s association with other disease outcomes, including Metabolic Dysfunction-Associated Steatotic Liver Disease (MASLD, previously termed NAFLD) and alcohol-induced liver steatosis, which we identified in our UKB analysis. Our *Liver fat (Retention)* cluster also had a small overlap (*PNPLA3* and *MAU2/TM6SF2*, ALT and AST) with the *Lipodystrophy 2* cluster in Smith et al. (2024)(23) - also derived from bNMF clustering, but applied only to T2D variants. Unlike the variants in our cluster that are all strongly associated with liver fat, their *Lipodystrophy 2* cluster included other variants and traits with no known association with liver fat. This could be why their *Lipodystrophy 2* PRS (polygenic risk score) (aligned with T2D-increasing alleles) showed an increase in CAD risk. If examined at the variant level, the literature has consistently shown a protective effect of variants in the *PNPLA3* and *MAU2/TM6SF2* genes on CAD. The differences between our cluster and Smith et al. (2024)(23) illustrate the advantage of clustering shared variants when investigating shared pathways between two diseases using bNMF, as it can reveal more specific mechanisms underlying both diseases that may be overlooked when clustering variants for one disease and then trying to identify associations with its comorbidities. Previous studies also found that T2D-risk increasing variant alleles in *PNPLA3*, *MAU/TM6SF2* and *APOE* were associated with increased hepatic lipid retention and reduced plasma lipid levels (55–57,59,60,68). Our metabolite results further support this, as our *Liver fat (Retention)* cluster and *Cholesterol* cluster exhibit a classical cardioprotective profile, which we resolve to specific VLDL, LDL, and HDL subclasses changes by leveraging metabolite GWAS(38).

The pathway involving increased liver fat, coupled with increased T2D risk and a discordant effect on CAD risk, captured by our *Liver fat (Retention)* and *Cholesterol* cluster, is also supported by other studies that focused specifically on the effects of liver fat. For example, Jamialahmadi et al. (2024)(68) generated two hypothesis-driven PRS for MASLD based on the presence of lipoprotein retention in the liver and found a distinct subtype of MASLD characterised by hepatic lipoprotein retention, increased risk of T2D and decreased risk of atherosclerosis. Ahmed et al. (2024)(57) identified 13 variants associated with liver fat using MRI data in the UKB and identified a cluster of variants involving *PNPLA3*, *TM6SF2*, *APOE* and *SUGP1* that showed a positive relationship between liver fat and T2D and an inverse relationship with CAD using two-sample MR analysis. Both studies link the group of liver fat (increasing) variants which were associated with increases in triglycerides and higher LDL to an increased risk of CAD. Our MR results between liver fat, T2D and CAD provide further support for the causal relationship between increased liver fat and the discordant risk of T2D and CAD. Through further analysis using partitioned MR, we showed that while liver fat was causally associated with T2D, the effect on CAD was driven by ApoB-containing lipoproteins rather than through liver fat directly. We summarise the proposed mechanism in **Figure 6**. Hepatic lipid retention leads to increased liver fat, and liver fat may be a causal mediator of T2D via hepatic fat-induced insulin resistance which reduces the inhibition of gluconeogenesis by insulin, resulting in fasting and postprandial hyperglycaemia while lipid synthesis is still upregulated via insulin regulated signalling pathways(69)(70). This can further worsen hepatic steatosis and leads to a self-reinforcing cycle. On the other hand, hepatic lipid retention decreases the level of plasma lipids, which reduces CAD risk independently of the increased T2D risk. The independent effect of these variants on decreased CAD is supported in individual-level analyses that identified a consistent effect on CAD after adjusting for T2D status in PRS analyses(24). Reduced circulating LDL and triglycerides have also been found in some studies to be associated with the risk of T2D(71,72), therefore there may be a second pathway that contributes to T2D via reduced plasma lipids due to hepatic lipid retention. These results may indicate that managing liver steatosis could be beneficial to T2D. However, if a patient has liver steatosis primarily due to hepatic lipid retention from carrying *PNPLA3/TM2SF6* variants, the risk of CAD may require careful monitoring, as reverting lipoprotein retention could increase serum triglyceride secretion. There are currently no approved medications for liver steatosis as the primary indication. Semaglutide, a GLP-1 agonist, which is indicated for T2D and has cardiovascular benefits(73–75) recently gained accelerated approval from the FDA to treat metabolic dysfunction-associated steatohepatitis (MASH). However, whether GLP-1s have beneficial effects in *PNPLA3/TM6SF2* genetic risk carriers is currently an active area of research(76,77). Therapeutic agents targeting *PNPLA3* are in active development, and early-phase trials have shown encouraging results for liver fat reduction with an acceptable safety and tolerability profile(78,79). It will be interesting to see if these therapeutics reduce the risk of T2D in secondary endpoint analyses; however, our findings suggest cardiovascular outcomes deserve explicit attention in those trials, as reversing hepatic liver retention could unmask ApoB-driven CAD risk in carriers.

**Figure 6.**
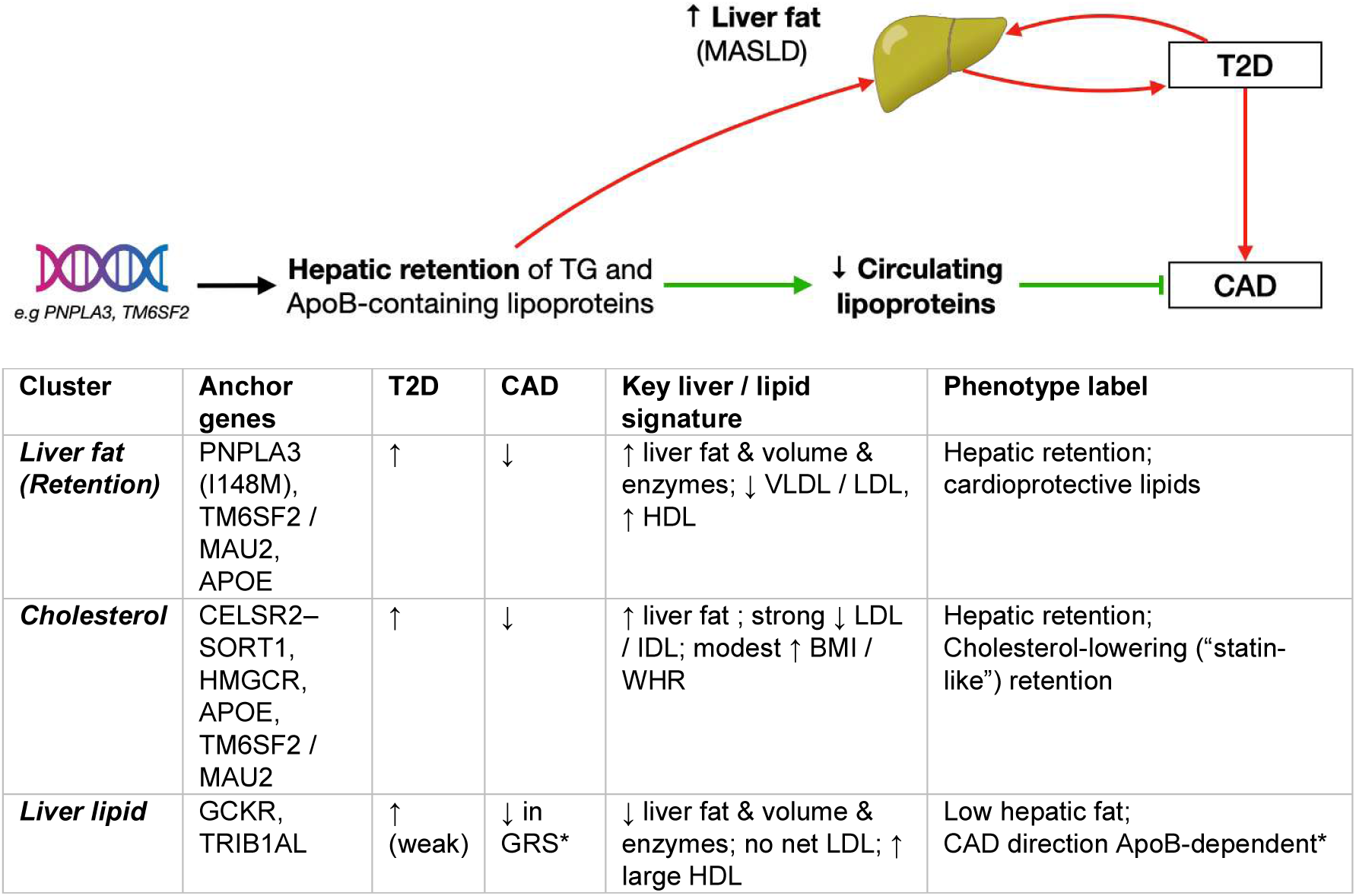
Summary of mechanisms linking increased liver fat with T2D and CAD. The schematic figure illustrates that not all liver fat carries the same cardiovascular meaning: whether it lowers or raises CAD risk depends on its effect on circulating lipoproteins =. The table summarises the three clusters that captured the discordant genetic risk between T2D and CAD involving liver fat, based on our clustering, metabolic traits analysis, UKB and MR for analyses. *In the ApoB-stratified MR, liver-fat variants that raise ApoB increased CAD risk; that grouping overlaps only partly with the Liver lipid cluster.

Our study has several strengths. First, we leveraged the statistical power of multi-ancestry T2D and CAD GWAS to identify multiple previously unreported shared signals, particularly discordant loci between T2D and CAD. Second, to our knowledge, we are the first to explicitly apply variant clustering approaches to a multimorbidity context by using shared variants. We showed that using shared variants yielded finer clusters and revealed additional biological insights specific to two diseases that were missed when clustering only variants associated with a single disease. Third, we leveraged recent large-scale GWAS of metabolomic traits and linked health record data to define diseases/conditions phenome-wide in the UKB, identifying many novel and distinct association patterns with our variant clusters that have not been described in other studies. Lastly, we use these data to elucidate further the relationships among liver fat, T2D, and CAD, providing insight into their connections and potential implications for management.

Our study has some technical limitations due to the methods used and the available data. We were limited in the colocalisation analysis by the lack of a multi-ancestry LD reference panel. We used a European reference panel, which was likely appropriate given the high proportion of European-ancestry individuals in the source GWAS. Our sensitivity analysis showed high consistency in colocalisation results when using European-only versus multi-ancestry summary statistics; thus, the risk of false positives due to differences in LD was low. Secondly, based on the availability of data, our analyses focused on data from primarily European-ancestry participants. Future work should investigate ancestry-specific variation, as there may be differences across ancestries(23).

In conclusion, we provided a comprehensive analysis of the genetic determinants and the concordant and discordant mechanisms shared between T2D and CAD. We showed that liver fat is a causal mediator for T2D while the effect on CAD depends on how the liver handles lipids (hepatic retention leads to decreased CAD risk and impaired export increases CAD risk) rather than from liver fat directly. We support monitoring cardiovascular outcomes in therapeutic agents reversing hepatic liver retention and the use of precision care to stratify disease/comorbidity prevention or treatment for T2D and CAD patients based on their specific underlying mechanisms.

## Data Availability

This research has been conducted using the UK Biobank Resource under Application Number 60847. Data from UK Biobank is available to registered researchers via application (https://www.ukbiobank.ac.uk/use-our-data/apply-for-access/).

## Acknowledgements

This research has been conducted using the UK Biobank Resource under Application Number 60847. This work uses data provided by patients and collected by the NHS as part of their care and support, ethics were approved by the North West Multi-centre Research Ethics Committee, we thank the participants for their valuable contributions to health data research.

XJ was supported by a Wellcome Trust and Health Data Research UK (HDR-UK) PhD studentship. NH is supported by a non-clinical PhD Studentship funded by AstraZeneca. EJN is supported by a Royal Commission for the Exhibition of 1851 Research Fellowship. SCR is supported by the British Heart Foundation Cambridge Centre of Research Excellence (CRE) Career Development Fellowship (RE/24/130011). This work was also supported by core funding from the British Heart Foundation (RG/18/13/33946; RG/F/23/110103), NIHR Cambridge Biomedical Research Centre (NIHR203312) [*], BHF Chair Award (CH/12/2/29428), Cambridge BHF Centre of Research Excellence (RE/18/1/34212), and by HDR-UK, which is funded by the UK Medical Research Council, Engineering and Physical Sciences Research Council, Economic and Social Research Council, Department of Health and Social Care (England), Chief Scientist Office of the Scottish Government Health and Social Care Directorates, Health and Social Care Research and Development Division (Welsh Government), Public Health Agency (Northern Ireland), British Heart Foundation and the Wellcome Trust. This work was performed using resources provided by the Cambridge Service for Data-Driven Discovery (CSD3) operated by the University of Cambridge Research Computing Service (www.csd3.cam.ac.uk), provided by Dell EMC and Intel using Tier-2 funding from the Engineering and Physical Sciences Research Council (capital grant EP/P020259/1), and DiRAC funding from the Science and Technology Facilities Council (www.dirac.ac.uk). *The views expressed are those of the authors and not necessarily those of the NIHR or the Department of Health and Social Care.

## Disclosures

A.S.B. reports institutional grants outside of this work from AstraZeneca, Bayer, Biogen, BioMarin, Bioverativ, Novartis, Regeneron and Sanofi. These companies had no involvement in the work presented here.

